# M&M: An RNA-seq based Pan-Cancer Classifier for Pediatric Tumors

**DOI:** 10.1101/2024.06.06.24308366

**Authors:** Fleur S.A. Wallis, John L. Baker-Hernandez, Marc van Tuil, Claudia van Hamersveld, Marco J. Koudijs, Eugène T.P. Verwiel, Alex Janse, Laura S. Hiemcke-Jiwa, Ronald R. de Krijger, Mariëtte E.G. Kranendonk, Marijn A. Vermeulen, Pieter Wesseling, Uta E. Flucke, Valérie de Haas, Maaike Luesink, Eelco W. Hoving, H. Josef Vormoor, Max M. van Noesel, Jayne Y. Hehir-Kwa, Bastiaan B.J. Tops, Patrick Kemmeren, Lennart A. Kester

## Abstract

With many rare tumor types, acquiring the correct diagnosis is a challenging but crucial process in pediatric oncology. Here, we present M&M, a pan-cancer ensemble-based machine learning algorithm tailored towards inclusion of rare tumor types. The RNA-seq based algorithm can classify 52 different tumor types (precision *∼*99%, recall *∼*80%), plus the underlying 96 tumor subtypes (precision *∼*96%, recall *∼*70%). For low-confidence classifications, a comparable precision is achieved when including the three highest-scoring labels. M&M’s pan-cancer setup allows for easy clinical implementation, requiring only one classifier for all incoming diagnostic samples, including samples from different tumor stages and treatment statuses. Simultaneously, its performance is comparable to existing tumor- and tissue-specific classifiers. The introduction of an extensive pan-cancer classifier in diagnostics has the potential to increase diagnostic accuracy for many pediatric cancer cases, thereby contributing towards optimal patient survival and quality of life.

## Introduction

Childhood cancer is the leading cause of disease-related deaths among children in high-income countries ^1^. A correct diagnosis is of the utmost importance to cure as many children as possible and improve quality of life ^2^. However, diagnosing tumors is not a trivial task with the plethora of tumors children can suffer from. While childhood cancer is rare in itself, within the collection of tumor entities there are many infrequently occurring ones, affecting less than one in 400 children. Diagnosis of rare tumor types tends to be more difficult due to their low frequencies and accompanying unfamiliarity to pathologists and hematologists, hereafter referred to as diagnostic specialists. Therefore, these tumors are associated with increased interobserver variability and above-average misclassification rates ^3–6^. To improve treatment selection and ultimately influence patient outcomes, the classification of especially rare tumor types needs to be more accurate.

One way to improve the diagnostic procedure is to use machine learning algorithms in healthcare ^7^. By performing computational classification, diagnostic specialists are provided with extra information that can either confirm their diagnosis or push the diagnostic process in the right direction. A methylation-profile based central nervous system (CNS) classifier, developed within the DKFZ in Heidelberg ^3^, was shown to increase patient survival ^8^. In recent years, several pediatric classifiers were developed, based on DNA-methylation or RNA-seq data ^3,9,10^. Nevertheless, these classifiers only cover a subset of the existing tumor (sub)types within pediatric cancer. The latest published pediatric tumor classifier, OTTER, was the first to utilize a pan-cancer approach ^11^. However, there are no direct links between OTTER’s classes and the diagnoses included in the WHO classification of pediatric tumors, making it less user-friendly for diagnostic specialists. Furthermore, OTTER can again only classify a subset of pediatric tumors. As a result, there is an urgent need for a more inclusive pan-cancer classifier to propel diagnostic utility.

Within the Netherlands, most children with cancer are diagnosed and treated in a single national pediatric oncology center where whole-exome sequencing and RNA-sequencing are routinely performed for diagnostic purposes. Of the available data, RNA-seq gene expression reflects behavior of tumor cells most directly, deeming this data informative for classifier development. Here, we present the Minority and Majority classifier (M&M), an ensemble-based RNA-seq machine learning approach to classify (rare) pediatric tumor entities in a pan-cancer setup. M&M can classify 52 different tumor types, together with their underlying 96 morphologically and/or biologically distinct tumor subtypes. As the machine learning algorithm is based on data sequentially collected for routine diagnostics in a national center, it has a more accurate representation of the pediatric tumor population than previous classifiers. For included tumor types, M&M reaches a diagnostic precision around 99%. To balance the classification accuracy across tumors with varying prevalence, we use an approach of integrating two classifiers with methods specifically focused on classifying rare tumor (sub)types (Minority classifier), or more frequent tumor (sub)types (Majority classifier). As a result, tumor types across the whole frequency range can be classified with comparable precision. Taken together, M&M has the potential to positively influence the diagnostic accuracy for pediatric tumors, likely resulting in increased overall survival and enhanced quality of life for children with cancer.

## Results

### Reference cohort and independent test cohort

RNA-seq data was collected from fresh-frozen samples within the Princess Máxima Center (PMC), the national Dutch pediatric oncology research hospital, for 3.5 years to establish a pan-cancer reference cohort. The cohort entailed primary, recurrent and metastatic tumor samples, samples before and after treatment, and non-neoplastic tissue samples. All samples were labeled according to the tumor domain, tumor type and tumor subtype they belong to (Figure 1, Extended Data Fig.1, 2). These labels were derived from the most recent WHO classification and provided by diagnostic specialists based on an integrated histomolecular approach. Computational classification was performed on the tumor type and subtype level. A minimum of three cases per tumor subtype was chosen to include as many rare pediatric tumor entities as possible. The resulting reference cohort contains a total of 1256 samples. As expected, the collection of the diagnostic population of patients resulted in a highly heterogeneous and imbalanced cohort, ranging from three to 269 samples per tumor type. In total, 52 tumor types, 96 underlying tumor subtypes and five non-neoplastic tissues were included (Supplementary Table 1). Pediatric diagnostic samples collected in the year following the reference cohort were used as an independent test cohort. In total, 471 samples from 442 patients were included, representing 39 tumor types, 70 tumor subtypes and 4 non-neoplastic tissues.

### Machine learning framework to classify rare tumor types

Within machine learning, there tends to be a bias towards the classes that contain most patients - the ‘majority’ classes. Consequently, recognition of infrequently occurring tumor (sub)types within a heterogeneous collection of tumor entities is particularly difficult. To reduce this majority-class bias, we created two different classifiers, a Minority classifier tailored towards correct classification of rare tumor (sub)types, and a Majority classifier with more predictive power for frequently occurring tumors. Each classifier was created using the same four steps of feature selection, feature reduction, down-sampling, and classification. These steps took place in different orders with different methods within the separate classifiers, to impose the specified classification focus (Figure 2a; Methods). Classification took place on the tumor subtype level, from which the tumor type could be extrapolated.

Extracting discriminatory information for each class is one of the principal steps within the development of the Minority classifier (Figure 2a, left panel). The F-statistic of analysis of variance (ANOVA) ^12^ was chosen to determine which transcripts differed most between tumor subtypes within the training set. The most variable transcripts were selected as input features and subjected to further reduction by determining each feature’s importance within a weighted Random Forest (RF) ^13^. To minimize class imbalance, the dataset with selected features was down-sampled 100 times to a maximum of three samples per tumor subtype. Each training subset was used for model development with weighted RF, generating a total of 100 models. Each model provided one classification label per sample, which was summarized to per-label probability scores.

The Majority classifier development (Figure 2a, right panel) was initiated with down-sampling. Again, 100 down-sampled training subsets were generated with now a maximum of 50 samples per tumor subtype. Feature selection and feature reduction took place within the subsets. The most variable transcripts across all data points were selected as input features for dimensionality reduction by principal component analysis (PCA). A weighted *k*-nearest neighbors (*kNN*) algorithm was used for model generation ^14^. The final classifications with their accompanying probability scores were determined using the same approach as for the Minority classifier.

After generating Minority and Majority classifications, tu-mor subtype labels were converted to their overarching tumor types for each classifier separately. The Minority and Majority classifier showed distinct performance characteristics across the pediatric tumor landscape (Extended Data Fig.3a). Frequently encountered tumor (sub)types displayed a better accuracy within the highest ranked classification labels, reaching saturation from 20 samples onwards (Figure 2b,c). The Majority classifier outperformed the Minority classifier for the top-ranked classifications across all tumor frequencies. Nevertheless, important to note is that for classifications with lower probability scores it is custom to look at more than just the top classification label. When taking into consideration the three highest-ranked classification labels instead, the correct label for rare tumor (sub)type classifications was included more often in the Minority classifier than the Majority classifier (Figure 2b,c).

**Fig. 1.**
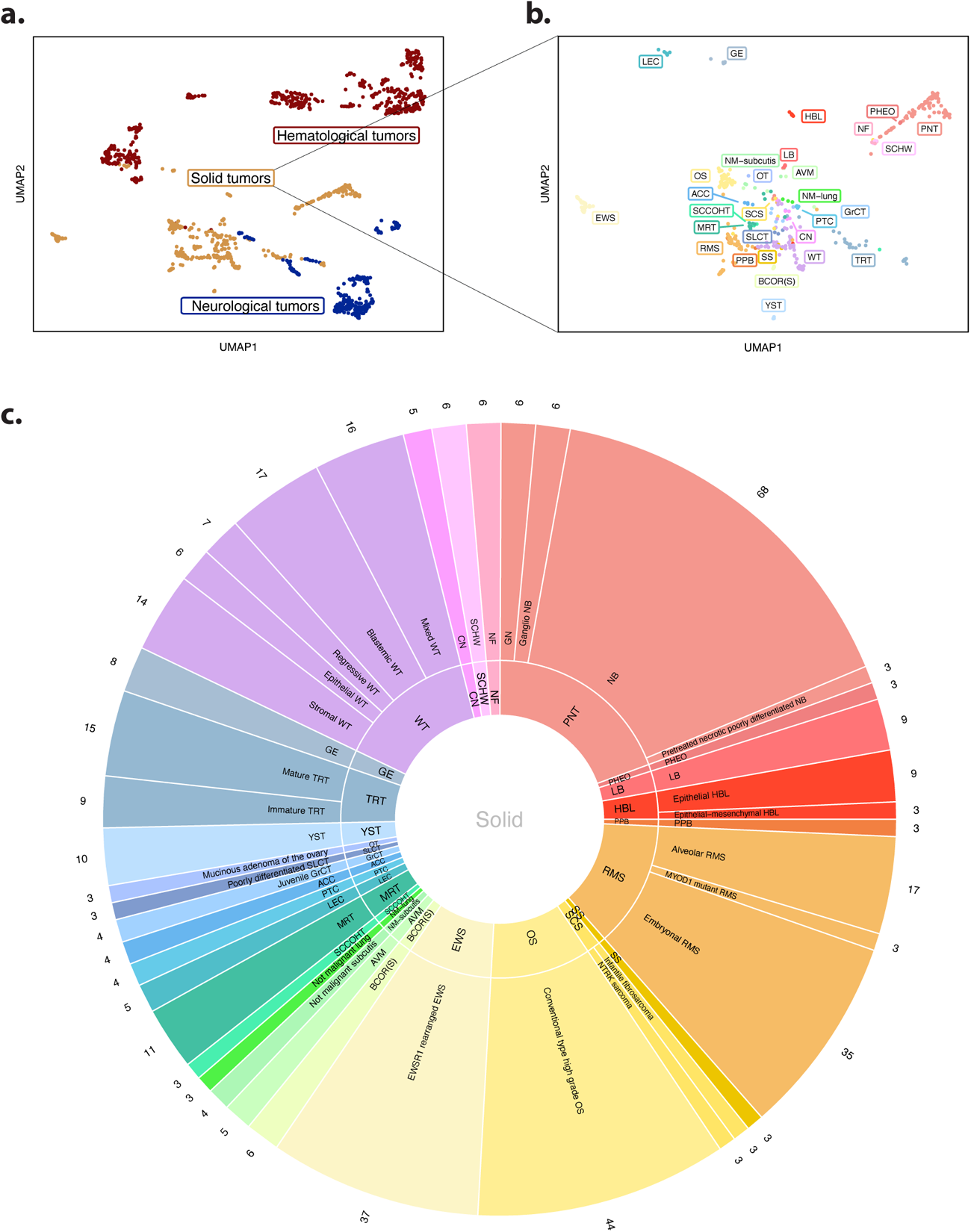
Overview reference cohort. **a)** Unsupervised clustering of reference cohort using UMAP, color-coded for the tumor domain the sample belongs to. **b)** Close-up of the UMAP projection of the solid tumor domain samples. Color coding is based on the underlying tumor type. Associations between abbreviations and tumor type names are available in Supplementary Table 1. **c)** Pie-chart showing the distribution of samples per tumor type (inner ring) and tumor subtype (outer ring) from the solid domain. Close-ups and pie-charts of samples from the hematological and neurological domains are shown in Extended Data Fig.1&2.

**Fig. 2.**
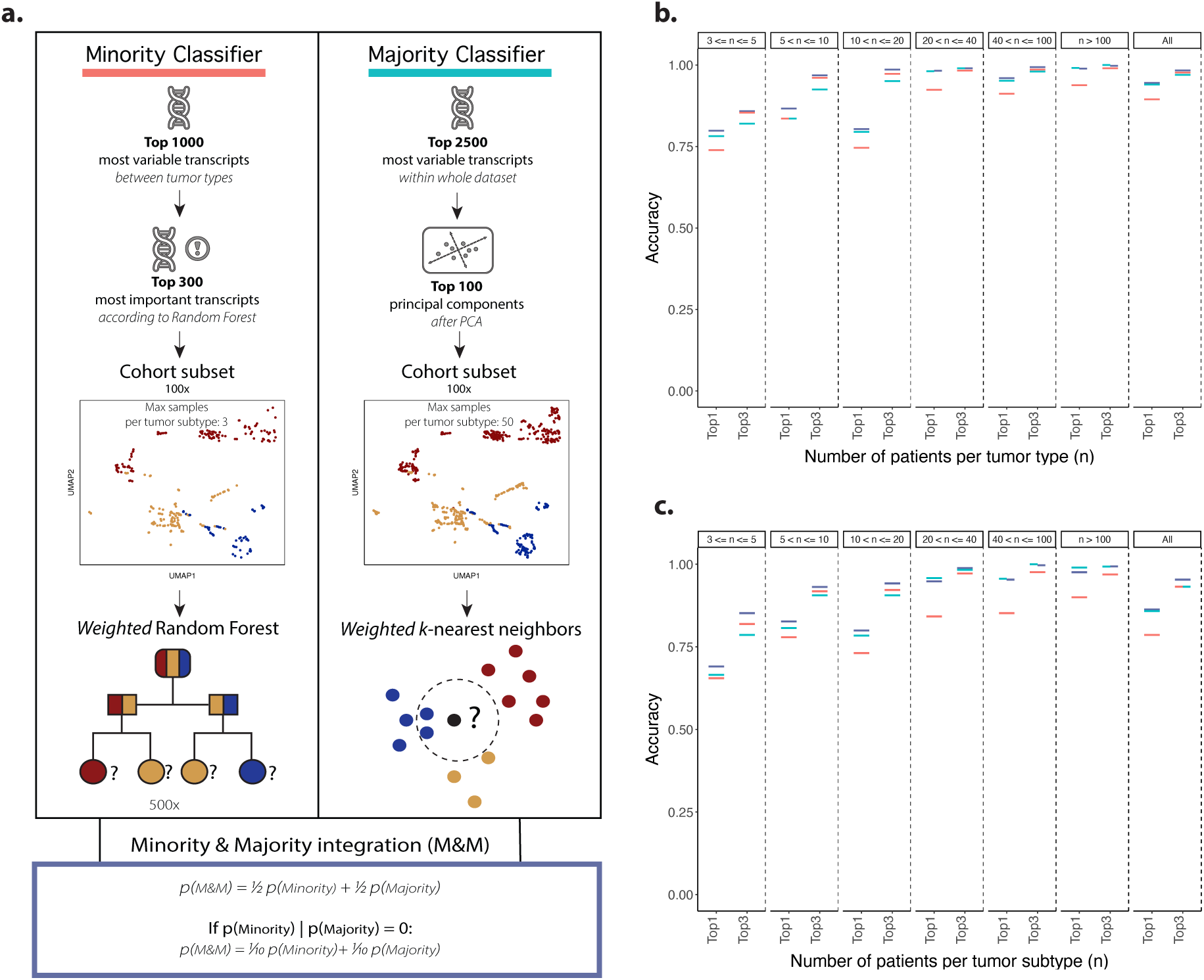
M&M framework. **a)** Schematic overview of the M&M framework, showing the separate Minority (left panel) & Majority classifier (right panel) machine learning workflows concerning feature selection, feature reduction, their down-sampling procedure, and their respective choice of algorithm. Note: the steps within the Majority classifier are not depicted in order, as cohort sub-setting takes place before feature selection. Classifier integration takes place after running the separate classifiers. The final probabilities were calculated by taking the average probability from the individual classifiers. If only one of the classifiers made a certain call, the final probability was divided by ten instead of averaged to penalize the classification label. **b,c)** Accuracy of separate Minority (red), Majority (blue), and integrated M&M classifiers (purple) for the tumor types **(b)** and subtypes **(c)** for different sample frequencies, determined in a ten-fold stratified cross-validation within the reference cohort.

Integrated classification label probability scores were calculated on both the tumor type and subtype level from the individual classifier probability scores (Figure 2a), as a measure of the confidence of M&M in the correctness of the classification. Recalibration of the probability scores could not be performed due to the inclusion of tumor (sub)types with low sample numbers (n*<*5). Ensemble-based classification resulted in more accurate calls for most tumors, preferentially boosting rare classes (Figure 2b,c, Extended Data Fig.3b). To balance sensitivity and specificity, high-confidence and low-confidence classifications were discriminated using a probability score cutoff determined by ROC-analysis (Extended Data Fig.4). Tumor subtype classifications were considered high-confidence if their probability score was higher than 0.72, while for tumor type classifications the probability score needed to exceed 0.82. In the end, M&M provides the three highest-scoring classification labels, hereafter referred to as top 3 classifications, with their associated probability scores (Supplementary Table 2,3).

**Fig. 3.**
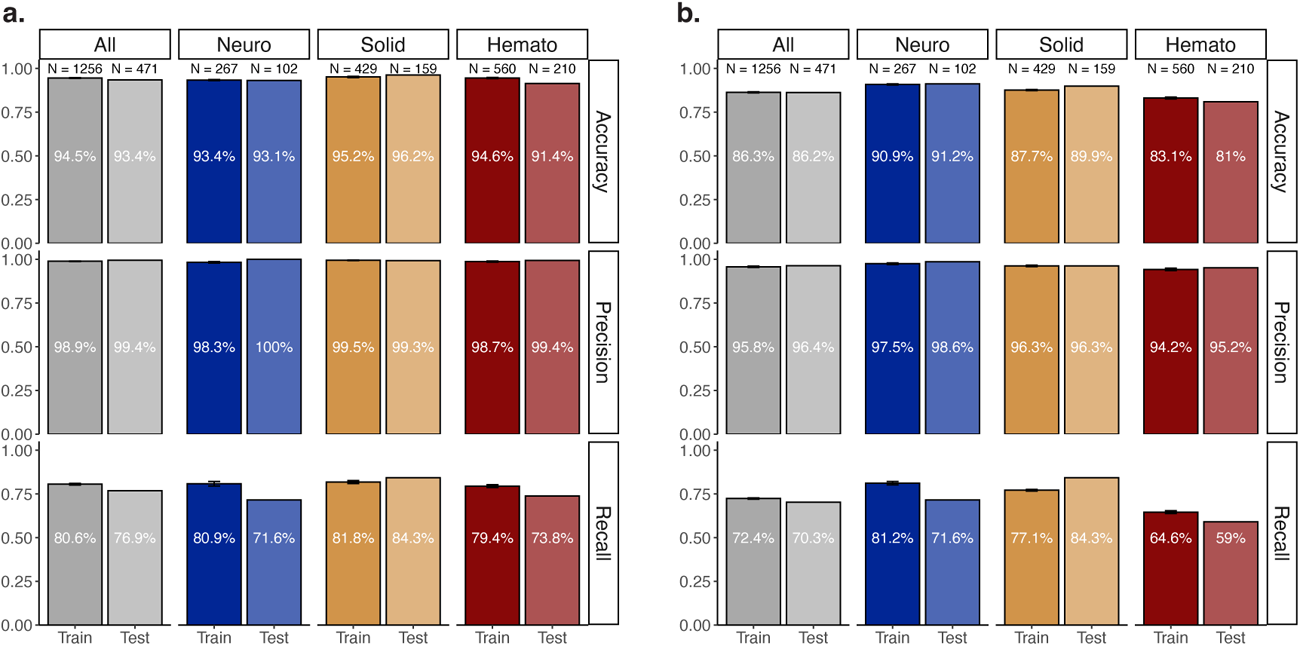
Performance M&M stratified by domain. Mean accuracy, precision and recall for the tumor type **(a)** and subtype **(b)** classifications for all samples (gray), the neurological (dark blue), solid (orange) and hematological domains (red). Results are shown for the reference cohort (dark bar, Train) and test cohort (light bar, Test). Error bars show the standard deviation for the performance measures achieved during ten independent runs of M&M.

A ten-fold stratified cross-validation was used on the reference cohort to estimate the efficacy of distinguishing different tumor (sub)types within M&M. The Minority and Majority classifiers were each run ten times, to gauge algorithm robustness. A single averaged accuracy, precision and recall were calculated from the different runs, together with their standard deviations. Recall was defined as the fraction of samples that were considered high-confidence classifications, with precision essentially being the accuracy within confident classifications. Average F1 scores, as the harmonic mean between precision and sensitivity, were calculated for each tumor (sub)type separately, together with their standard deviation. Furthermore, a single, final classification label per sample was obtained by averaging classification probabilities per individual label over the ten runs and selecting the highest scoring one for each sample.

### Cohort-wide performance M&M

M&M’s main aim is to be clinically implemented within a pediatric oncology setting. Therefore, the method’s outcomes were focused on diagnostically relevant properties: the performance for confidently classified samples and associated recall, and the performance for the low-confidence classifications within the top 3 labels. M&M could correctly classify the tumor type for 94.5% of the samples within the reference cohort (Figure 3a), and the underlying tumor subtype for 86.3% (Figure 3b). For high-confidence tumor type classifications, M&M could reach a precision of 98.9% for *∼*80% of the samples (N = 1003/1256 samples). On the tumor sub-type level, a precision of approximately 96% was achieved for *∼*70% of the samples (N = 911/1256 samples). Slight differences in results were observed between the domains, with solid tumors showing the highest overall precision and recall. On the subtype level, neurological tumors were classified with the highest accuracy, precision and recall. Given that precision scores for all three domains were above 98% for tumor types and 94% for subtypes, M&M portrayed a remarkable performance for included classes across the pediatric tumor landscape.

**Fig. 4.**
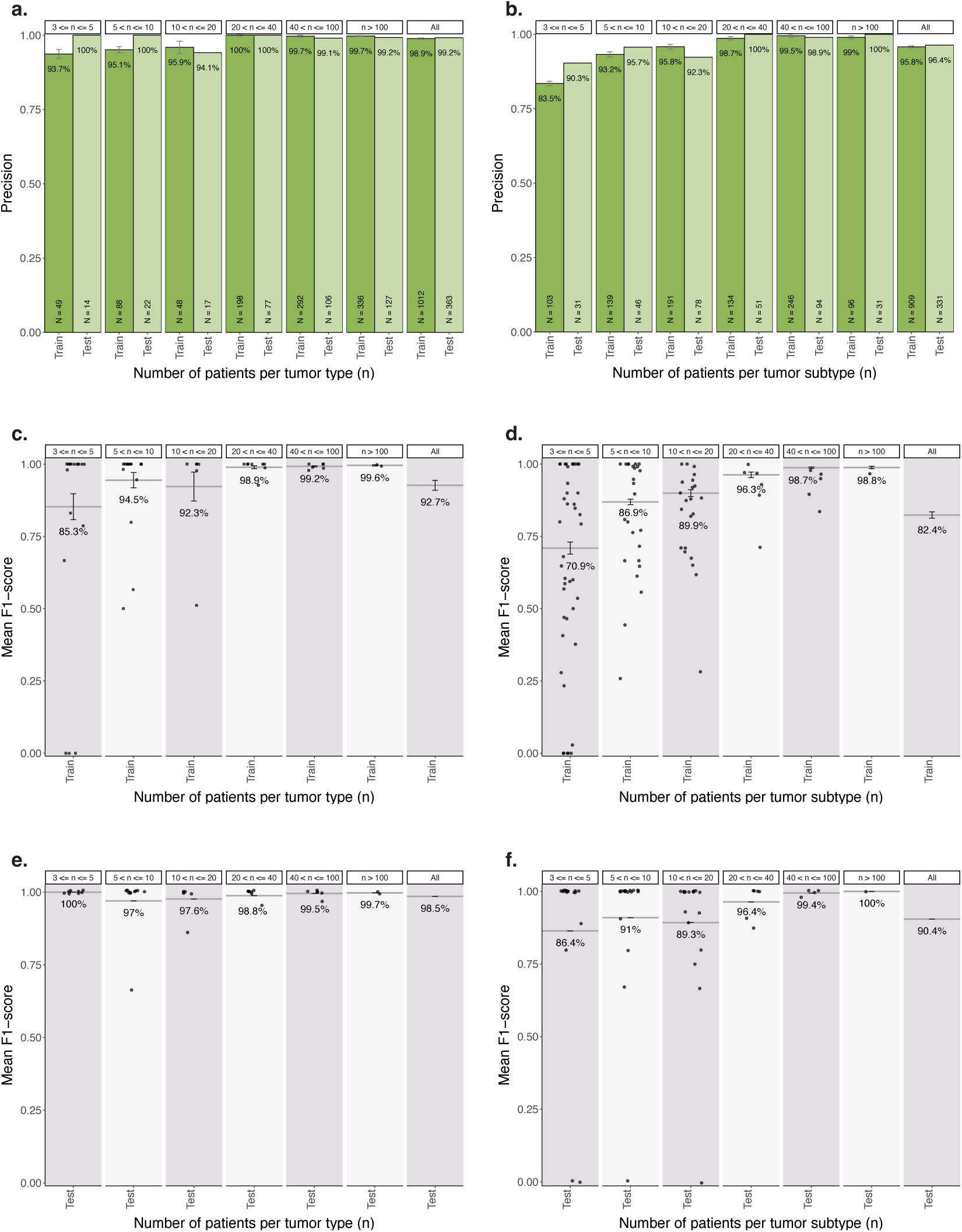
Performance M&M on separate tumor entities, stratified by tumor (sub)type frequency. **a,b)** Precision of M&M for the high-confidence tumor type **(a)** and subtype **(b)** classifications for different sample frequencies within the reference cohort (dark bar, Train) or test cohort (light bar, Test). **c-f)** Average F1 scores for the tumor type **(c,e)** and subtype **(d,f)** classifications for different sample frequencies within the reference cohort **(c,d)** and test cohort **(e,f)**. F1-scores of individual tumor entities are displayed by jittered black dots.

**Fig. 5.**
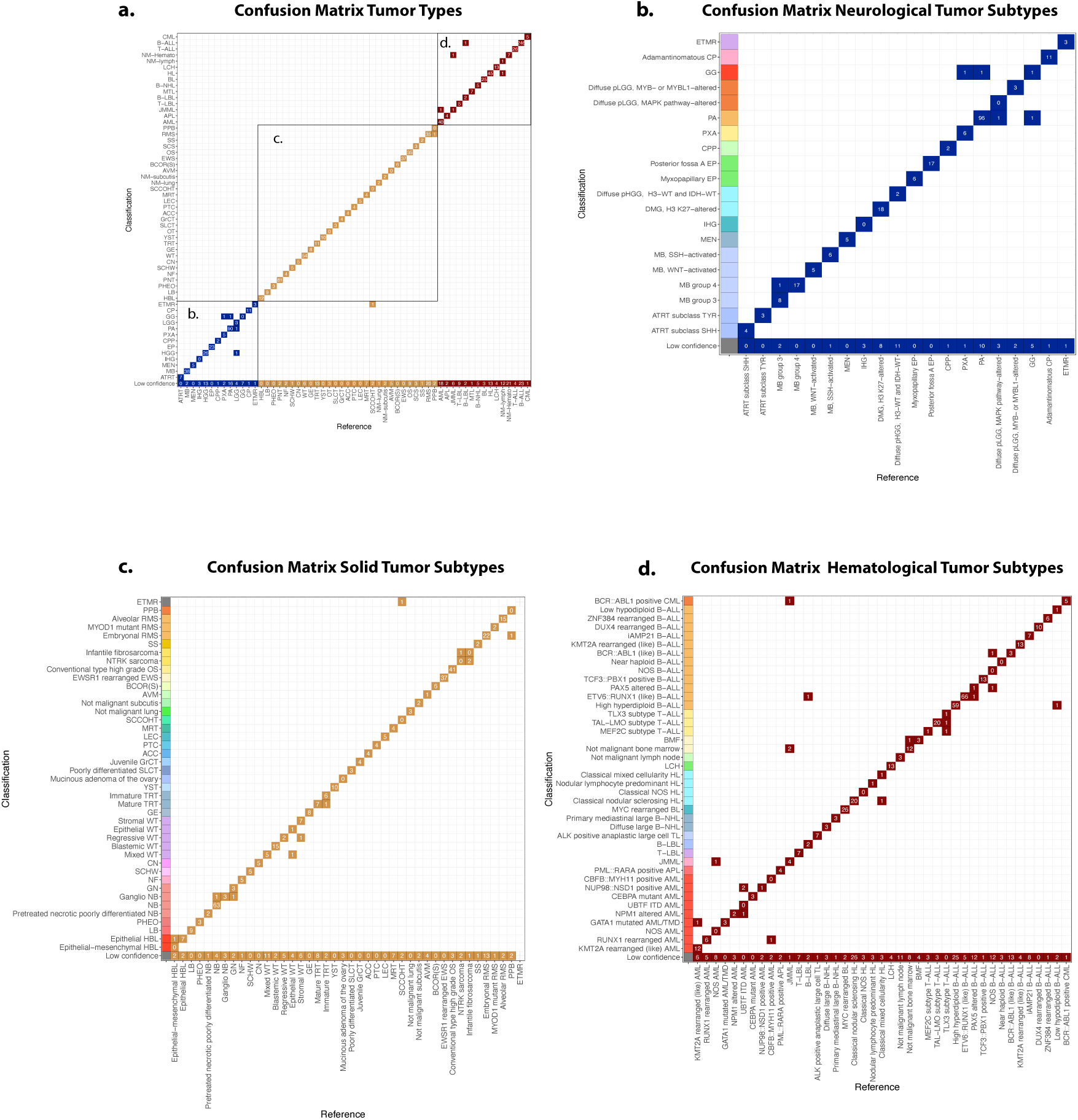
Misclassifications between different tumor entities. Confusion matrix of all tumor type **(a)** and subtype **(b-d)** predictions from the neurological **(b)**, solid **(c)** and hematological **(d)** domains, within a 10x cross-validation setup. The gold standard (Reference), provided by diagnostic specialists, is specified on the x-axis, while M&M’s call (Classification) is located on the y-axis. Within the blocks, it is specified how often a certain classification is made. Furthermore, it is portrayed how often a tumor type or subtype does not reach the confidence score threshold (’Low confidence’). Color-coding on the left within **b-d** is used to show which tumor subtypes belong to the same overarching tumor group.

An independent test cohort was used to validate the results obtained for the reference cohort, specifically testing for potential overfitting on the training data. First, it needed to be checked whether M&M could identify samples from patients diagnosed with tumor subtypes not present within the reference cohort, by providing low-confidence classifications. Out of the 580 samples that passed quality control measures, 109 samples (19%) from 98 unique patients represented tumor subtypes that were not covered within the reference cohort. 92 of these samples (80%) did not obtain a high-confidence classification on the tumor subtype level, showing M&M’s capacity to recognize when samples deviate from all available tumors within the algorithm. Of the 17 remaining samples, 9 were confidently classified as a different tumor subtype within the same parent tumor type (8%), while 4 were classified based on the tissue signal (4%). All samples without a matching diagnosis in M&M were removed from further analyses, resulting in a test cohort of 471 samples. Here, M&M could achieve a tumor type classification precision of 99.2%, which was even higher than the 98.9% obtained for the reference cohort (Figure 3a). Tumor subtype classifications showed a comparable test cohort precision of 96.1% versus 95.8% for the reference cohort (Figure 3b). Again, similar performances were observed across the different domains, together demonstrating M&M’s robustness in performance.

**Fig. 6.**
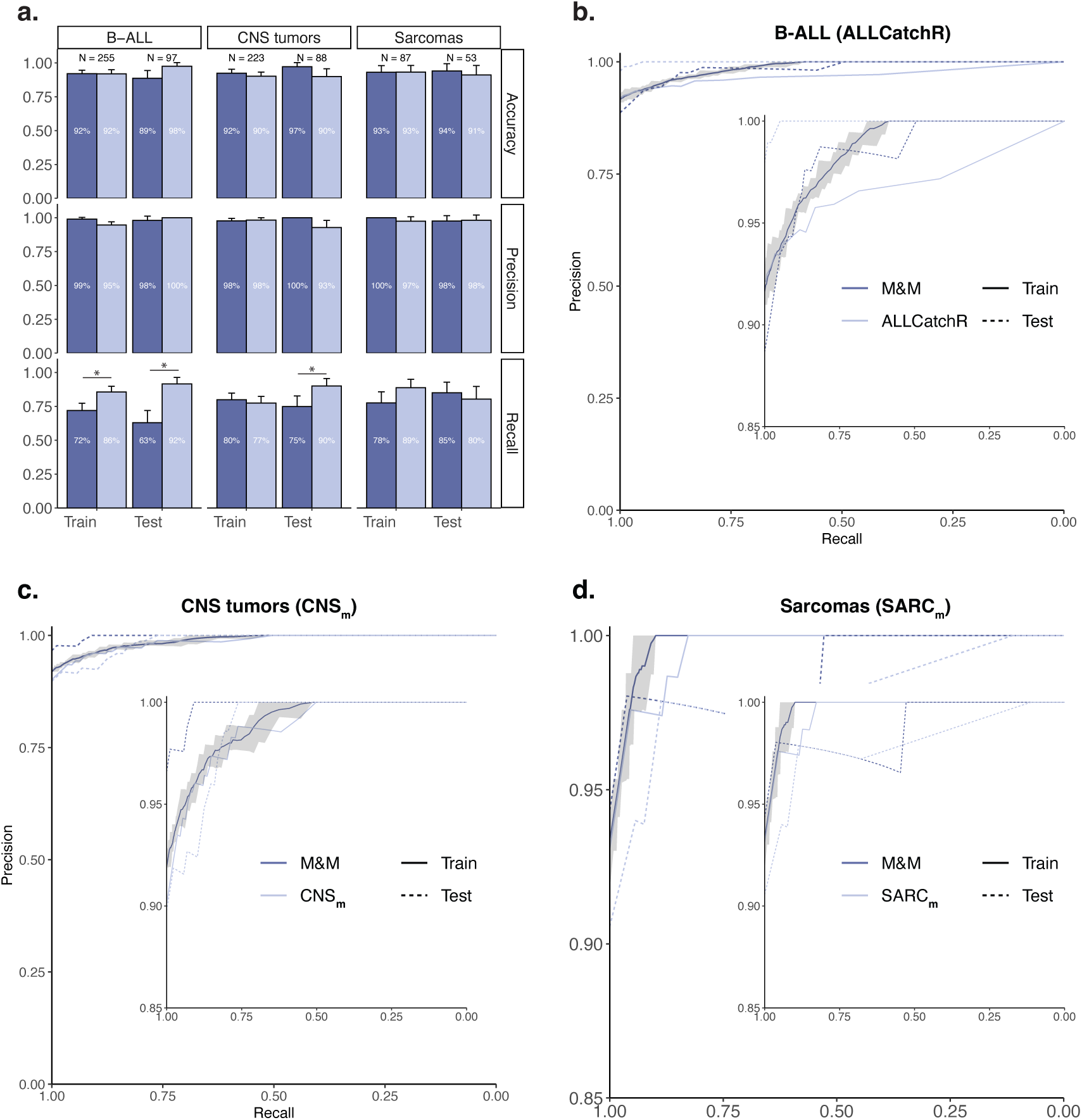
Performance of M&M compared to more class-restricted tumor classifiers. **a)** Barplot visualizing the average accuracy, precision and recall of M&M (dark color) compared to the other classifiers (light color) - ALLCatchR (left), the DKFZ central nervous system methylation classifier (CNS*m*, middle) and the DKFZ sarcoma classifier (SARC*m*, right) - on the corresponding tumor samples from the reference cohort and test cohort, subsampled 100 times to 25% of the available samples to generate different subsets. Error bars represent standard deviations. A star (*) signifies a significant difference (p-value *<* 0.05). **b-d)** Precision-recall (PR) plots for the comparison between M&M (dark color) and the other classifiers (light color) - ALLCatchR **(b)**, CNS*m* **(c)**, and SARC*m* **(d)** - for the reference cohort (solid line) and test cohort (dashed line). The shadow indicates the lower and upper precision score at a specific recall rate observed within ten independent cross-validation runs of M&M on the reference cohort. A close-up of the precision range of 0.85 to 1 for the PR plot is displayed within each PR curve.

For low-confidence classifications, it was of interest how often M&M could provide relevant tumor (sub)type classifications within the top-ranking labels. The latter is essential to guide diagnostic specialists during the diagnostic process for samples that have a less clear-cut diagnosis. Within the top 3 labels, the correct tumor type diagnosis was present in 98% of the cases, in both the reference and test cohort (Extended Data Fig.5a). For tumor subtypes, approximately 95% of the reference and test cohort samples could be assigned a correct label within the top 3 classifications (Extended Data Fig.5b). These numbers were only slightly lower than the precision scores for high-confidence classifications, showing that for low-confidence classifications the correct tumor (sub)type label is generally present within the top 3 labels.

### Rare tumor types can be classified with high precision

To enable some generalization of classification results, tumor types and subtypes were grouped according to their respective frequency within the reference cohort. Based on these subsets, it appeared that both rare and frequently occurring tumor types within the pediatric population were accurately classified by M&M (Figure 4a), resulting in a minimum precision of 93.7% for rare tumors (3-5 samples), and 99% for tumor types with over 100 samples. Recall rates were lower for rare tumor types (Extended Data Fig.6), however, still 68% of samples were classified confidently. M&M portrayed a high average F1 score (*∼*0.93) for tumor type classifications (Figure 4c,e), indicating that individual tumor types can be accurately classified and labels are discriminatory between classes. Rare tumor types portrayed lower average F1 scores. However, this result appears highly influenced by a few extreme outliers, while most minority class tumor types still achieved perfect classification. Combined, these results show M&M’s strength for rare tumor type classification.

As expected, tumor subtype classifications showed lower overall precision and F1 scores compared to tumor type classifications, with low-frequency subtypes showing the lowest scores (Figure 4b,d,f). Specifically, multiple subtypes with 3-5 samples in the reference cohort appeared ill-defined by M&M (F1 *<* 0.25). Nevertheless, these rare subtypes again displayed low recall rates, resulting in a final precision of approximately 84%.

### M&M misclassifications are infrequent, non-repetitive and independent of sample type

Only a limited number of tumor type misclassifications occurred within the high-confidence classifications of the reference cohort (Figure 5a, Supplementary Table 4). Most misclassifications occurred once and were confined within their domain of origin. In total, one cross-domain error happened between two primitive embryonal tumors. Tumor subtype misclassifications occurred at higher frequencies within overarching tumor types in all three domains (Figure 5b-d). Furthermore, 88% of samples from subtypes that had a ‘not otherwise specified’ (NOS) label were misclassified as other entities within the same tumor type. This indicates that the label’s non-specific nature might negatively influence their classification accuracy. In general, the tumor subtype mix-ups would not have resulted in a change of treatment regimen for the patient, and hence most errors would not have had direct clinical consequences.

M&M was run on samples stemming from primary, recurrent and metastatic tumors. Furthermore, some patients received systemic chemotherapy before tumor sample acquisition, while others did not. While analyzing M&M’s performance on separate sample types, the precision on tumor type level was highest for primary tumors and untreated samples (*∼*99%) (Extended Data Fig.7). This result was expected, as these samples most accurately reflect the underlying tumor biology. Nevertheless, the difference in precision with recurrences, metastases and systemically treated samples was max 3%, with approximately equal fractions of samples receiving high-confidence classifications. To get some insights into the influence of the sequencing protocol and RNA-seq quality on M&M’s performance, eight formalin-fixed and paraffin-embedded (FFPE) tissues with available RNA-seq data were classified as well. M&M could classify seven out of eight samples correctly (Extended Data Fig.7), with the incorrect classification being low-confidence. Therefore, these preliminary results indicate that M&M may be capable of classifying both fresh-frozen and FFPE samples.

### M&M performs comparable to existing, more class-restricted pediatric tumor classifiers

When introducing a pan-cancer classifier, it is important to check whether it can compete with other classification algorithms without causing a performance drop. To this end, we compared M&M’s results to three well-known and well-performing pediatric tumor classifiers: ALLCatchR for B-ALL tumor subtypes, the DKFZ methylation classifier for CNS tumor types (CNS*_m_*) and the DKFZ methylation classifier for sarcomas (SARC*_m_*) ^3,9,10^. To make the comparison robust, 100 different data subsets were generated by down-sampling to 25% of the available samples. Subsequently, an average accuracy, precision and recall were determined from the performance within the subsets, together with their associated standard deviation. M&M portrayed comparable accuracy and precision scores (Figure 6a, Supplementary Table 5-7). However, M&M obtained more low-confidence classifications, resulting in a significantly decreased recall rate in half of the comparisons (p *<* 0.05). The test cohort performance measures were comparable to the findings in the reference cohort, providing more confidence in observed findings.

To further compare the classifiers’ performance, precision-recall (PR) curves were generated. For M&M, the ten classifier runs were combined to generate one minimum, average and maximum precision at each recall score (Figure 6b,c,d). M&M appeared to slightly outperform ALLCatchR for the B-ALL sample classifications within the reference cohort, while in the test cohort the roles were reversed (Figure 6b). The seemingly large difference in performance on the test cohort between ALLCatchR and M&M could be explained by similarities in expression profiles between tumor subtypes *BCR::ABL1* (like) B-ALL, iAMP21 B-ALL and *PAX5*-altered B-ALL ^15,16^. Test samples had comparable probability scores for the labels within M&M. Nevertheless, the correct label was present for all but one sample within the three highest scoring classification labels (Supplementary Table 4), underscoring the importance of looking at more than just the top-scoring classification. For the methylation-based classifiers, M&M showed comparable PR-curves within the reference cohort (Figure 6c,d). Within the test cohorts, M&M had higher PR-curves at the highest recall rates, indicative of enhanced classification accuracy. Taken together, M&M can compete with established domain or disease-specific classifiers on included classes, thereby showing its potential as a pan-cancer classification algorithm.

## Discussion

There is a large variety of tumor types a child can suffer from, making it difficult at times for diagnostic specialists to provide the correct diagnosis. Unfortunately, an incorrect diagnosis can be detrimental to the child’s odds of survival and quality of life, as it may lead to an incorrect or suboptimal treatment regimen. To improve the diagnostic procedure for pediatric tumors, we developed an ensemble-based pan-cancer classifier, M&M, able to classify tumors based on RNA-seq expression profiles. To maximize the diagnostic utility, M&M includes as many rare tumor (sub)types as possible given the currently available data.

Both tumor type and subtype classifications, defined by clinicopathological and histological tumor characteristics, could be accomplished with a performance comparable to other classifiers that are focused on a subset of pediatric cancers. RNA-seq based classification by M&M showed similar performance to both methylation-based classifiers it was compared to, despite having been trained on a considerably smaller dataset. Also, M&M’s comparable accuracy to the Heidelberg CNS methylation classifier is remarkable as current WHO labels for neurological tumors are in part based on the outcome of this classifier. M&M’s suitability for tumor subtype classification is endorsed by its comparable performance to ALLCatchR, an algorithm specifically designed for B-ALL subtype classifications only. Most importantly, besides the samples that are captured within these existing classifiers, M&M covers 29 additional pediatric tumor types and 55 subtypes within the solid (N = 29) and hematological domain (N = 26), showing its immediate added benefit for pediatric tumor classification.

M&M was developed with the goal to strive towards an even performance, regardless of tumor (sub)type. By using a novel way of limiting the influence of highly abundant tumor (sub)types, M&M could achieve a generally balanced accuracy across different sample frequencies. Rare tumor types with as little as three patient samples could be included within the reference cohort, resulting in a classification precision of 99% for all samples and 94% for rare tumor types. Most tumor types were recognized near-perfectly within the datasets, while samples of less well-classifying tumor types were mostly classified as low-confidence. As expected, tumor subtype classifications showed worse performance than tumor type classifications. However, the drop in precision for high-confidence classifications was, with 3%, modest. The additional tumor subtype misclassifications resulted mainly from rare tumor subtypes classifying as a more frequent subtype within the overarching parent tumor type. This phenomenon suggests more samples may be needed for subtype than for tumor type discrimination.

To our knowledge, M&M is the most comprehensive pediatric tumor-specific classifier so far, spanning the entire domain of pediatric tumors. Furthermore, M&M encompasses samples from all tumor stages, treatment statuses and from several non-neoplastic tissues. The algorithm is capable of working with ribo-depleted fresh-frozen tissue, and some initial results suggest that RNA-seq from FFPE tissues can also be analyzed. Within the PMC, this possibility extends the clinical usability to essentially all obtained patient samples. M&M’s workflow greatly simplifies the user experience, only requiring RNA-sequencing data and one classifier. Furthermore, the algorithm classifies a new clinical RNA-seq sample in less than five minutes, with algorithm updating requiring approximately an hour. Additional verification showing the prospective performance in an extended clinical setting is needed for standard-of-care use, both in the current center but also when implementing the algorithm in other centers. Amongst others, it should be checked more thoroughly whether FFPE samples can be classified reliably, and whether it would be possible to analyze poly(A)-enriched RNA-samples as opposed to ribosomal RNA-depleted samples with M&M as well.

Important to note is that M&M is intended to assist specialists during the diagnostic procedure, not as a stand-alone replacement for the diagnostic procedure. M&M can now directly provide insights into the molecular make-up of the tumor to indicate which disease the patient is suffering from, so that subsequent relevant molecular tests can be performed. This agnostic setup is especially powerful for tumors that are hard to confirm via classical histopathology. For example, a sarcoma with a BCOR genetic alteration can be readily identified by M&M with high confidence for all samples (Figure 5), while classical histopathology has difficulty distinguishing it from other small blue round tumors.

The M&M algorithm showed similar performance statistics for primary, recurrent and metastatic tumor samples, suggesting that classification accuracy was mostly independent of tumor stage. Especially for metastatic tumors, this is a striking result, showing that tumor signal within RNA-seq data can be recognized independent of the surrounding tissue. This idea is further supported by the notion that M&M can correctly discriminate between several non-neoplastic and neoplastic lesions. As an example, osteosarcoma metastases, often residing in the lungs, are never classified as non-neoplastic lung tissue. Consequently, the chance that M&M is merely a tissue-classifier is deemed minimal. Further sources of sample heterogeneity, such as tumor treatment status and even FFPE, did not appear to have a large impact on the classification accuracy, suggesting that differences in sample types might not necessarily have a large influence on M&M’s performance.

For some tumors, M&M had difficulty separating them from each other. Inadequate feature selection might result in seemingly equal transcriptomes within different tumor entities. Furthermore, classification errors could potentially portray that the RNA-sequencing data simply might not contain information for the separation of certain entities. Particularly, this could be the case with tumors whose labeling is partially driven by clinical information. For example, both spindle cell sarcoma (SCS) subtypes - infantile fibrosarcoma and *NTRK* sarcoma - have alterations in the *NTRK*-gene. The main difference is based on the age at diagnosis of the patient (infantile fibrosarcoma *<* 1, *NTRK* sarcoma *>* 1). Tumor type classification as SCS is perfect within samples from these classes, while the subtype distinction is mainly wrong (Figure 4). Alternatively, M&M could indicate the presence of multiple distinct molecular subgroups currently captured within one tumor label, leading to an incoherent signal. This phenomenon likely plays a role in the NOS misclassifications, where there may be a variety of currently unknown genetic events underlying the development of the tumor, each resulting in a distinct expression profile. M&M has the potential to pinpoint tumor entities which are currently not necessarily well-defined by their label and help further shape tumor class definitions.

An area of interest for future developments of M&M is the inclusion of even more rare tumor (sub)types. Especially for the neurological tumors, the inclusion of more (sub)types would enable the gradual replacement of CNS*_m_* by M&M, which would currently not be recommended due to missing subtypes. As there is a continuous flow of new incoming diagnostically used RNA-seq samples, samples of extremely rare tumor (sub)types will accumulate over time, eventually reaching the threshold for inclusion into M&M. When combining the samples from the reference cohort and test cohort, an additional 10 tumor types, 18 tumor subtypes and 3 non-neoplastic tissues could already be included within the classifier (Supplementary Table 8). Alternatively, the number of extremely rare tumor samples could potentially be increased in-silico, speeding up their inclusion.

A correct diagnosis within pediatric oncology becomes more and more important, as it is increasingly recognized that many (molecular) subtypes may benefit from emerging subtype-specific therapy. Complementing diagnostics with RNA-seq based classification has the potential to help with guiding therapeutic decision making, thereby allowing for an optimal treatment regimen for patients. Besides using RNA-seq for its expression data, it additionally contains information on the presence of fusion-genes, and/or other specific genetic alterations. This allows for an integrated analysis where it is possible to both suggest a tumor (sub)type, and confirm this by identifying the molecular driver (fusion/mutations) within the same diagnostic assay. In this way, M&M will likely contribute to a better future for pediatric oncology patients, not only for their survival, but also for an improved quality of life.

## Data Availability

All data produced are available or will be available online on Github, Zenodo and ArrayExpress.

https://github.com/princessmaximacenter/MnM

https://zenodo.org/records/11575098

## Methods

### Obtaining patient material and RNA-seq data

RNA-seq data was obtained from patients with a suspected neoplasm using the same methods as described by Hehir-Kwa et al. (2022) ^17^. In summary, either a tissue or liquid biopsy was taken for diagnostic purposes, or from tumor material obtained during resection. All tissues to be subjected to RNA-seq were fresh-frozen, with the exception of 8 samples that had previously been formalin-fixed and paraffin-embedded. Sequencing libraries were generated using 300 ng RNA. The ribodepletion protocol KAPA RNA HyperPrep Kit with RiboErase (Roche) was used, generating target insert sizes of approximately 300 base pairs. Sequencing was performed on a NovaSeq 6000 system (2x 150 base pairs, lIlumina). The resulting reads were aligned using Star fusion (version 2.7.0f) to GRCh38 and gencode version 29, and base qualities were recalibrated with GATK4. Quality control was performed using Fastqc (version 0.11.5) and Picard (version 2.20.1). Expression counts were eventually calculated at transcript level by Subread Counts. All types of RNA transcripts were included. Potential RNA-isoforms were not taken into consideration. Instead, one ‘meta-transcript’ was generated, combining counts of all exons within one transcript. In total, raw RNA-seq counts were gathered for 58804 transcripts. Subsequent normalization to transcripts per million (TPM) was performed.

### Cohort selection

Fresh-frozen tissue and liquid biopsy samples collected from 01-12-2018 until 01-06-2022, for which consent was obtained through the Princess Máxima Center for Pediatric Oncology biobank, were considered for inclusion into the reference cohort. Samples of primary, recurrent and metastatic tumors were all present in different proportions. Both samples from systemically treated and untreated tumors were included. Most, but not all systemically treated tumors were recurrent or metastatic tumors. Suspected neoplasms were sequenced as well, extending the cohort to include some non-neoplastic tissue samples. The dataset was as unbiased in nature as possible. However, participation bias could result from differences in the frequency of patients providing informed consent per tumor entity. Important to note is that retinoblastoma is absent from the reference cohort, as these patients are treated within the Amsterdam University Medical Centers (location VUmc) instead of the PMC.

For inclusion, the sample needed to be of a pediatric patient, with an age ranging from 0 to 18. Further inclusion criteria were sufficient RNA-seq quality as determined by a minimum of five million unique reads, less than 85% duplicate reads, and a sufficient tumor purity (*>*5% for all tumors except Hodgkin lymphoma (HL) and Burkitt lymphoma (BL); for those no minimum purity) as manually determined by a diagnostic specialist, and a maximum of one sample per patient per tumor subtype to guarantee a high-quality cohort. Samples that were suspected to be involved in sample swaps were removed. The labels of the samples were based on the most recent WHO classification ICD-O-derived codes; however, more specific labels than the current ICD-O-3 standard were used where relevant. Contrarily, certain labels were collapsed onto a more general class label, governed by the known absence of differences in expression profile. For example, patients that either suffered from B-ALL with a *BCR::ABL1* fusion, or B-ALL with an expression profile similar to the samples with the fusion (*BCR::ABL1* like B-ALL), were collapsed into the class *BCR::ABL1* (like) B-ALL. Labels were an integrated histomolecular diagnosis provided by diagnostic specialists, based on histology, immunohistochemistry, methylation array data and PCR-sequencing analysis. A lower limit of three samples per tumor subtype was chosen for inclusion in the machine-learning models.

Eventually, 1256 samples were selected within the reference cohort. Tumor stage and systemic treatment status was known for more than 96% of those samples, with mainly non-neoplastic samples not being assigned a status (n = 35 / 39). As an independent test cohort, all pediatric diagnostic samples collected between 01-06-2022 and 01-09-2023 were considered (N = 704). Samples from patients who were also included in the reference cohort were excluded to prevent bias (N = 87). Samples with unclear diagnosis, bad biopsy or RNA quality, or ones that were potentially involved in sample swaps were removed (N = 27). Furthermore, test samples with extremely low tumor purity (*<*5%, with the exception of HL and BL) were considered unrepresentative and unusable within computational classification, and thereby excluded (N = 10). Of the remaining samples, 109 had a tumor subtype diagnosis that was not represented within M&M (19%), so these samples were removed as well, resulting in a final test cohort of 471 samples. In addition to the test cohort, eight FFPE tissues were analyzed separately.

### RNA-seq preprocessing

Ribosomal RNA depletion (ribo-depletion) leaves a different number of residual rRNA-transcripts within the final RNA-seq count data per sample based on the protocol efficiency. To increase data consistency, a linear model was generated to predict what percentage of RNA-molecules was protein-coding. The 5000 transcripts with the highest mean expression within the reference cohort were selected as input features for the model. Subsequently, these features were normalized to be centered around zero, using the mean and standard deviation. The centered features were used as input features for a generalized linear model with elastic net regularization ^18^ to predict nonprotein-coding percentages. R package glmnet ^19^ (version 4.1.7) was used for model generation. Hyperparameter tuning of regularization parameter lambda was performed using a ten-fold cross-validation setup. Only features with a coefficient higher than 0.01 were selected for use within the final linear prediction model to prevent overfitting. Subsequently, the non-protein-coding fraction was predicted and converted to a predicted protein coding fraction. For each sample, transcript counts were divided by the predicted fraction of protein-coding molecules, increasing the counts proportionally to the amount of rRNA-pollution within the dataset. Furthermore, transcripts that anti-correlated substantially with the protein-coding fraction, as determined by the linear model coefficients, were removed from the dataset. In total, 58774 transcripts remained in the cleaned dataset, which were subsequently log_2_-transformed.

### Classifier cross-validation setup

Classification was performed on the tumor subtype level, from which the parent tumor type could be inferred. The reference cohort was split up into ten parts, in which stratification ensured approximate equal distributions of the tumor subtype samples over the parts. Subsequently, nine parts were used as training data, while the residual part was set aside and only used in the end to determine the generated classifier’s performance. To estimate the robustness of the generated classifier, the cross-validation process was repeated ten times, using different seeds that resulted mainly in different data stratification, different training data subsets and different trees within RF. After manual optimization of the workflow, one final classifier was generated to classify new samples, using all reference cohort samples as training data. Overfitting within this final classifier was evaluated by classifying samples of the independent test cohort.

### Minority-class focusing workflow

Within the Minority classifier workflow, a classical equal-variance one-way ANOVA is used to determine which transcripts differ most between groups, which can subsequently be selected as input features. The difference between groups is estimated by the F-statistic, a measure based on calculated group averages and calculated standard deviations per transcript. TPM counts were used instead of the log_2_-transformed counts as input values for ANOVA. Important to note is that ANOVA could only be performed if there were at least three samples per group. Unfortunately, for tumor (sub)types with three samples in total, only two samples remained within the training dataset if the test cohort contained one of the tumor samples. The issue was resolved by temporarily adding one synthetic data entry for tumor subtypes with only two available samples, which were again removed from the dataset after feature selection. The synthetic data entry was created by taking the average expression for all transcripts of these samples and using these as input values.

Downsampling was performed by subsampling the complete training dataset 100 times, generating 100 different training subsets in total. To this end, initial sampling with replacement was performed, keeping in the unique entries. The per-sample sampling probability was determined to be the inverse of the square root of the total number of tumor subtype samples 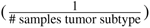 within the dataset, resulting in higher sampling probabilities for samples of lowly abundant tumor subtypes. A minimum of one sample per tumor subtype was imposed. Subsequently, downsampling of larger classes to three samples per tumor subtype was performed, resulting in training sets containing between one and three samples per tumor subtype.

A weighted RF was utilized both for further feature reduction and the final model generation, using the randomForest package (version 4.7.1.1) in R ^20^. No hyperparameter tuning was performed. Default settings included that the number of transcripts provided at each split was the square root of the total number of transcripts in the dataset, and random sampling with replacement was used to select training samples for each tree. In total, 500 binary decision trees were generated. The weighted setup of RF linked each tumor subtype to its prior, which is subsequently used during bootstrapping. The priors were chosen to be the reciprocal of the total number of samples per tumor subtype within the training set.

The total number of features was reduced to 300, using a feature importance score metric calculated within RF itself. The feature importance score was determined by calculating the mean decrease in accuracy upon removal of the feature from RF. The setup was run on the 100 training subsets, using the 1000 most variable transcripts selected via the F-statistic of ANOVA. The importance of the features in the 100 different training sets was averaged and ordered, after which the top 300 features were selected as input variables for the final model development. The same training subsets and weighted RFsetup were used to generate 100 different Minority models.

### Majority-class focusing workflow

The Majority classifier setup is initiated with the same downsampling procedure as performed for the Minority classifier, except now the maximum number of samples per tumor subtype is increased from three to fifty. Each training set eventually contained one to fifty samples per tumor subtype. Within each training subset, feature selection is performed by selecting the 2500 features that portray the highest variance. All features were centered around zero using the mean and standard deviation per transcript. PCA was performed to reduce dimensionality within the dataset further. A manual optimization procedure showed that the first hundred principal components (PCs) contained most information (data not shown), so that these features were kept for the final model generation. Weighted *kNN* from the R package kknn (version 1.3.1) ^21^ was used to create a model within each training data subset. Leave-one-out cross-validation was performed to determine the optimal number of neighbors, ranging from one to a maximum of 25 neighbors. The ‘optimal’ kernel, a Minkowski distance of 1 and no scaling were used. The weighted variant of *kNN* takes the distance from the test point to each of the k nearest neighbors into account, which is particularly useful in situations of class imbalance. Hyperparameter tuning was performed using a ten-fold cross-validation setup.

### Majority-voting

M&M makes use of a majority-voting system, where the different votes come from classifications provided by models that were generated on different training datasets. Eventual probability scores for a classification label are calculated as the fraction of models that called a sample the same label. To come to an ensemble-based classification, integration of the two individually generated scores is needed. The sum-rule for classifier probability score combination, considered the most restrictive and robust ^22,23^, was used to integrate the Minority and Majority classifier results into M&M. Equal weights were given to the Minority and Majority classifier scores. If supporting information for a classification label was absent from either one of the classifiers, resulting in a single-classifier probability score of zero, the classification label was not trusted. For these labels, the probability score was divided by ten instead of by two. The resulting probability scores cannot be interpreted as the real-life probability of correctness of a classification, due to the inability to perform model recalibration.

To estimate at which probability score there is an optimal balance between removing erroneous classifications (high specificity) and keeping in as many correct classifications as possible (high sensitivity), an ROC-analysis was performed. In this case, ‘positives’ were samples that remained classified, while ‘negatives’ were samples that did not reach the probability score threshold and were thereby removed. An optimum was defined as the probability score threshold at which the sensitivity and the specificity are balanced. For the tumor type classifications, this score was around 0.82 (Extended Data Fig.4a), while for the tumor subtype classifications the score was determined around 0.72 (Extended Data Fig.4b).

### Comparison to other classifiers

Only samples from tumor entities that could be classified by both classifiers, as specified within the classifier documentation, were selected, considering that each classifier had a specific area of expertise within the pan-cancer setting. ALLCatchR could be run on all B-ALL samples except the NOS B-ALL, with RNA-seq data availability being the only requirement. For the CNS and sarcoma classifiers, on the other hand, DNA methylation data was required. Of the 262 tumor samples from the neurological domain within the reference cohort, 224 could be analyzed and matched to an existing class using the CNS methylation classifier (85%). Out of 171 sarcoma samples within the reference cohort, 89 methylation profiles were available for analysis by the Sarcoma methylation classifier (*∼*52%). More data was available for the test cohort, having 88 CNS methylation samples out of the 98 CNS test samples (*∼*90%) and 53 sarcoma methylation samples out of the 66 sarcoma test samples (*∼*80%). After obtaining classification labels for each of the three other classifiers, label conversion was performed to match the corresponding tumor (sub)types labels with M&M’s labels.

For the CNS and sarcoma classifiers (CNS*_m_* and SARC*_m_*), version 12 and version 10 were used respectively. Samples with low probability scores (*<*0.3) obtained the classification label ‘Not classified’ within the methylation-based classifiers, resulting in these being counted as misclassifications. To minimize their influence on the PR-curves, the probability scores were manually put at 0, so that all these classifications were removed simultaneously at the earliest possible score. Furthermore, M&M’s classification labels with probability scores below 0.3 were also converted to ‘Not classified’, guaranteeing a fair comparison. As a result, M&M’s actual performance is often better than currently portrayed.

The resulting dataset was subsampled 100 times to approximately 25% of the available samples, using random sampling without replacement. Within each subset, the accuracy, precision and recall were calculated using probability score thresholds described within the original guiding articles. Specifically, only the high-confidence classifications were selected to calculate the precision on, while the recall was calculated as the fraction of high-confidence classifications within the whole subset. For CNS*_m_*and SARC*_m_*, confidence score thresholds of 0.84 were used ^3,9^. ALLCatchR’s results included information on which samples were called with high confidence, so that these could directly be inferred. Subsequently, the obtained performance measures for the 100 data subsets were compared by counting in how many subsets each classifier outperformed the other one. Resulting p-values evaluating the performance were calculated as 1 minus the fraction of subsets in which a classifier outperformed the other one. The performance was considered significantly enhanced when a classifier outperformed the other classifier in at least 95 out of the 100 subsets, as a proxy for a p-value equal to or below 0.05.

### Data and Software Availability

To facilitate M&M’s use on other datasets, an R-package for its implementation has been created, under the name of MnM. The package is freely available on Github (https://github.com/princessmaximacenter/MnM), while future efforts will try to implement it in CRAN. Included are options to retrain the separate Minority and Majority classifiers, check their performance within a ten-fold cross-validation setup, and use the newly trained M&M classifier for new sample classifications. To check classifier performance more extensively, options are implemented to further visualize the results. A tutorial is provided on Github to allow for easy implementation of the algorithm and visualization tools. The final trained Minority & Majority classifiers generated on the reference cohort will furthermore freely be available on Zenodo as R-objects, which can be used for new sample classifications using the MnM package. Important to note is that missing transcript imputation is currently not implemented, so all transcripts that are used as features need to be available within the dataset. The original transcript per million data of the reference cohort and test cohort used for classification purposes are available within ArrayExpress (accession E-MTAB-14038).

### Software and data visualization

The whole setup was performed with R software (version 4.2.3) ^24^. Data visualization was performed using the R packages ggplot2 (version 3.5.1) ^25^, umap (0.2.10.0), ungeviz (version 0.1.0) and ggrepel (version 0.9.5.9999). Figures were optimized using Adobe Illustrator 2023.

## Declarations

### Ethics and reporting

Pediatric patients diagnosed between December 2018 and September 2023 at the Princess Máxima Center for Pediatric Oncology, Utrecht, the Netherlands, were eligible upon the following criteria: (1) availability of RNA-sequencing (RNA-seq), (2) informed consent for the biobank study. No clinical exclusion criteria were applied. The study was approved by the Biobank and Data Access Committee (BDAC) of the Princess Máxima Center for Pediatric Oncology. All included patients provided written informed consent for participation in the biobank (International Clinical Trials Registry Platform, NL7744).

## Extended data

**Extended Data Fig. 1.**
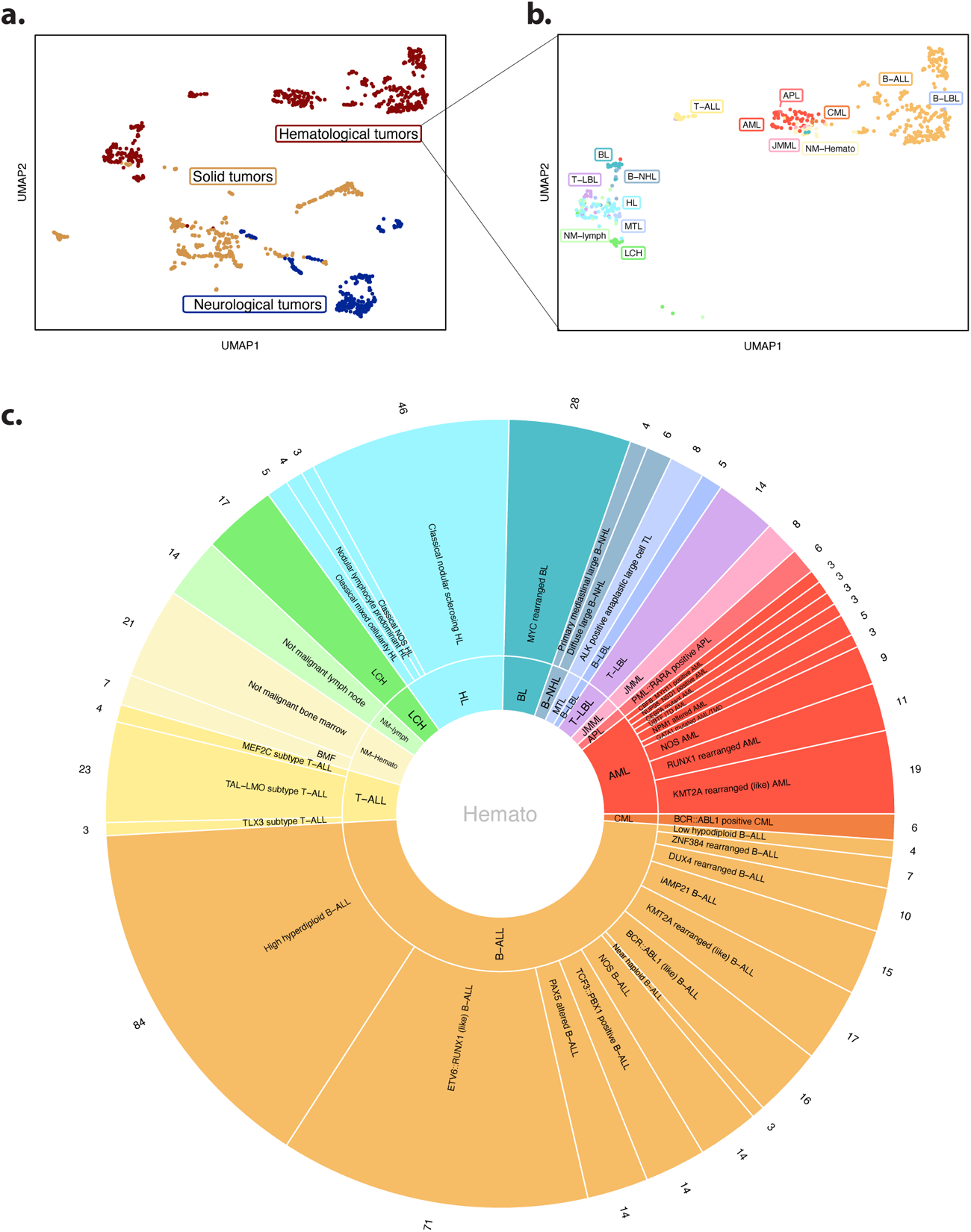
Overview reference cohort. **a)** Unsupervised clustering of reference cohort using UMAP, color-coded for the tumor domain the sample belongs to. **b)** Close-up of the UMAP projection of the hematological domain samples. Color-coding is based on the underlying tumor type. Associations between abbreviations and tumor type names are available in Supplementary Table 1. **c)** Pie-chart showing the distribution of samples per tumor type (inner ring) and tumor subtype (outer ring) from the hematological domain.

**Extended Data Fig. 2.**
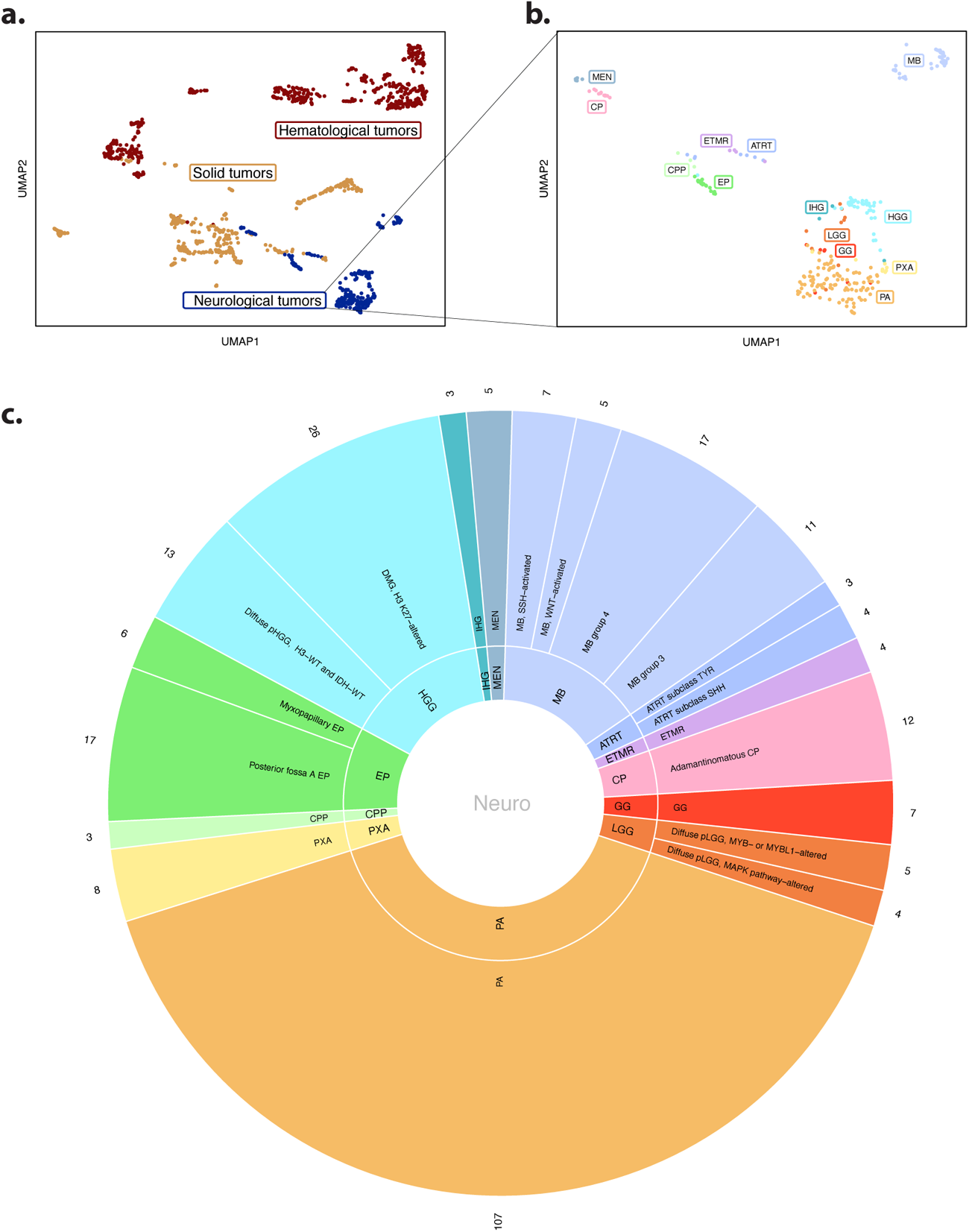
Overview reference cohort. **a)**: Unsupervised clustering of reference cohort using UMAP, color-coded for the tumor domain the sample belongs to. **b)** Close-up of the UMAP projection of the neurological domain samples. Color-coding is based on the underlying tumor type. Associations between abbreviations and tumor type names are available in Supplementary Table 1. **c)** Pie-chart showing the distribution of samples per tumor type (inner ring) and tumor subtype (uter ring) from the neurological domain.

**Extended Data Fig. 3.**
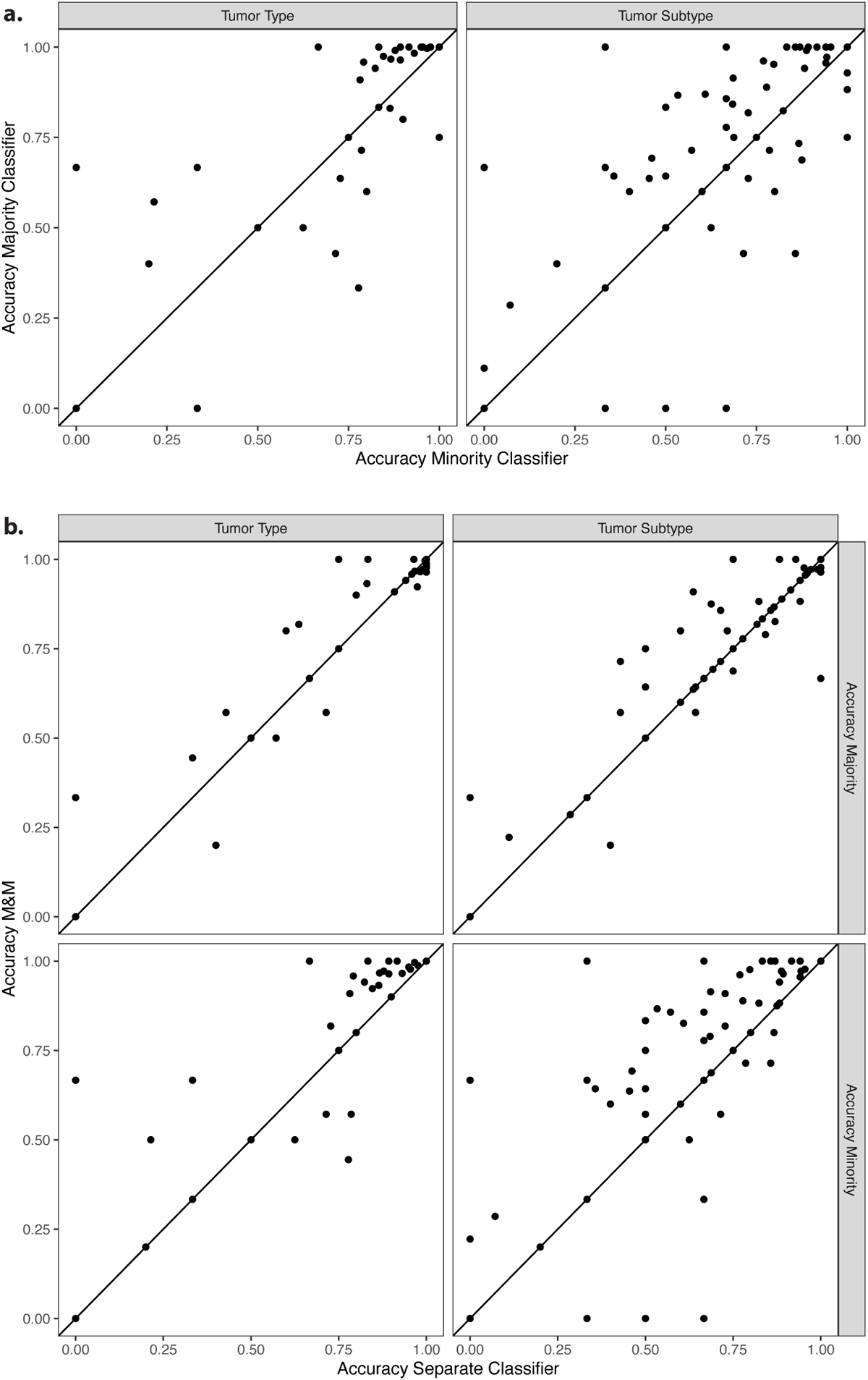
Accuracy of all classifiers on separate tumor (sub)type classifications. **a)**: Accuracy of the Minority (x-axis) and Majority Classifier (y-axis) on tumor types (left panel) or tumor subtypes (right panel) relative to one another. **b)** Accuracy of the separate Minority (x-axis, lower panels) and Majority classifier (x-axis, upper panels) compared to the accuracy of integrated classifier M&M (y-axis), on tumor types (left panels) and subtypes (right panels).

**Extended Data Fig. 4.**
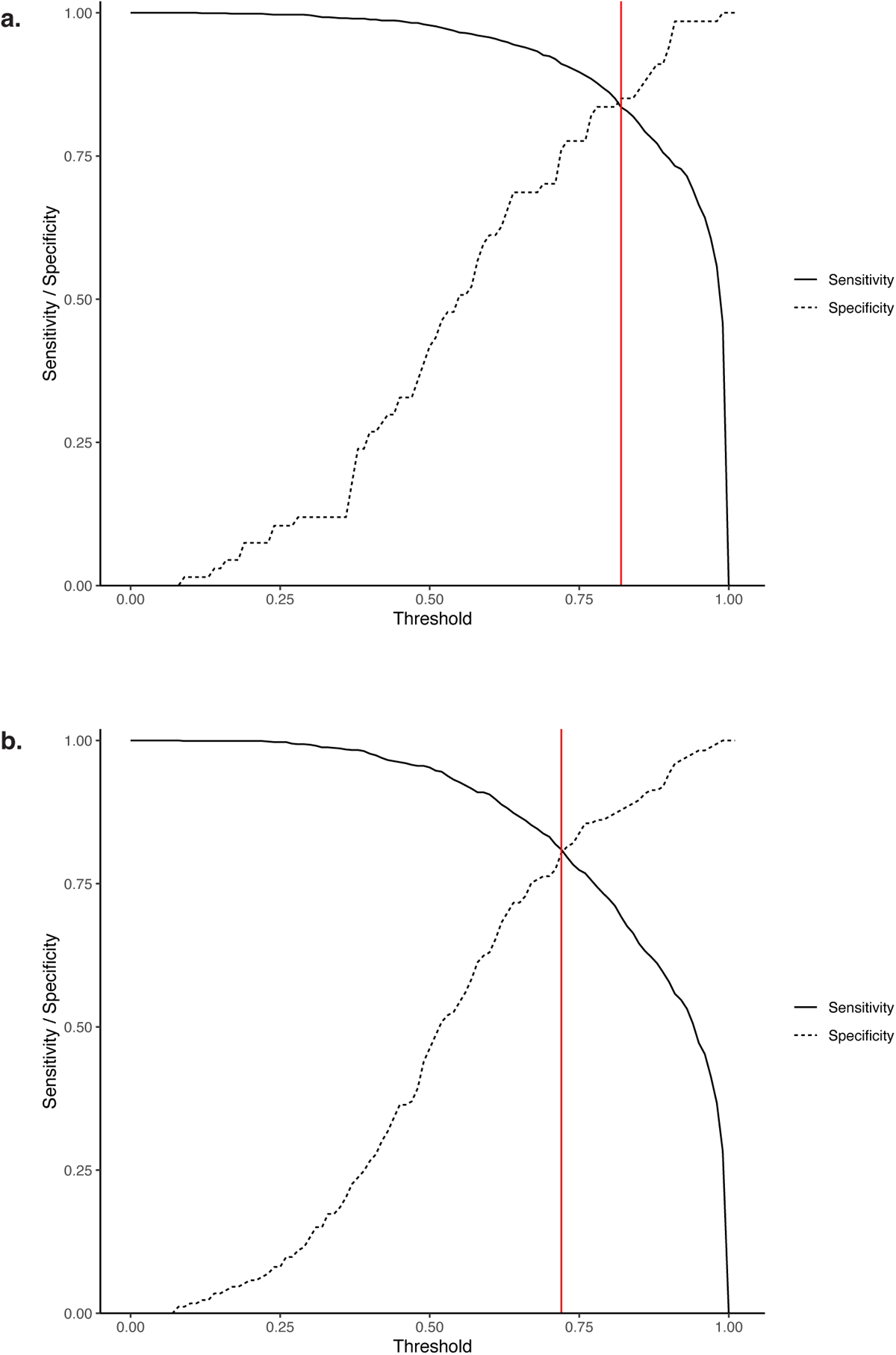
Determination of probability score threshold for separation of high-confidence and low-confidence classifications.: Receiver operating characteristic (ROC) curve analysis to determine at which threshold the optimal balance between the sensitivity and specificity was achieved. The red line shows the chosen threshold for the probability score for the tumor type **(a)** and tumor subtype **(b)**: 0.82 and 0.72.

**Extended Data Fig. 5.**
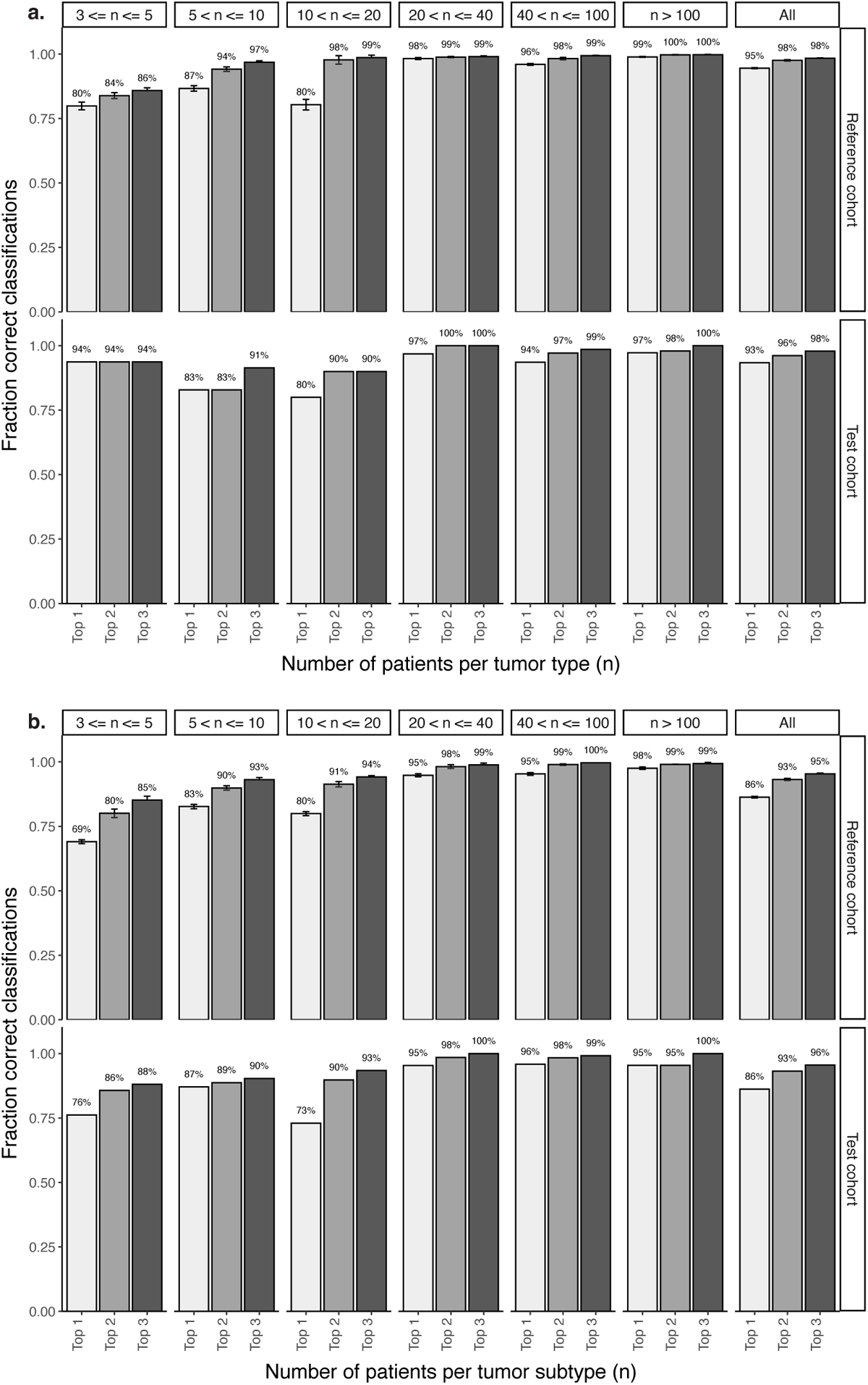
Percentage of correct diagnoses within M&M’s top 3 classification labels. **a,b)** Shown are the results for the complete set of tumor type **(a)** and subtype **(b)** classifications within the ten-fold cross-validation on the reference cohort (upper panel) and independent test cohort (lower panel).

**Extended Data Fig. 6.**
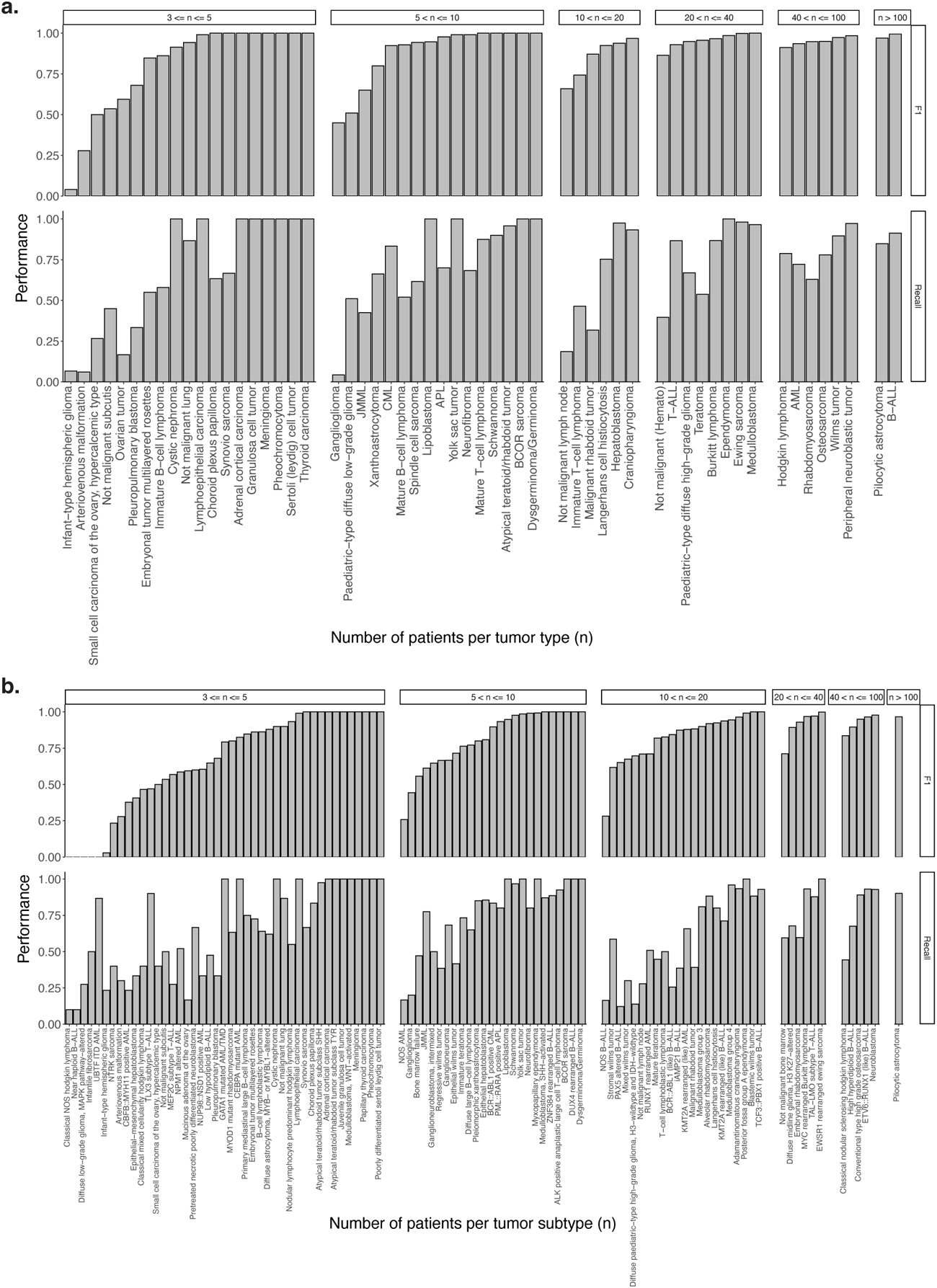
F1 score and recall per pediatric tumor entity, stratified by population frequency. a-b) It shows how often a tumor type (a) and subtype (b) is correctly classified within the reference cohort, compared to how often the probability score reaches the threshold (Recall).

**Extended Data Fig. 7.**
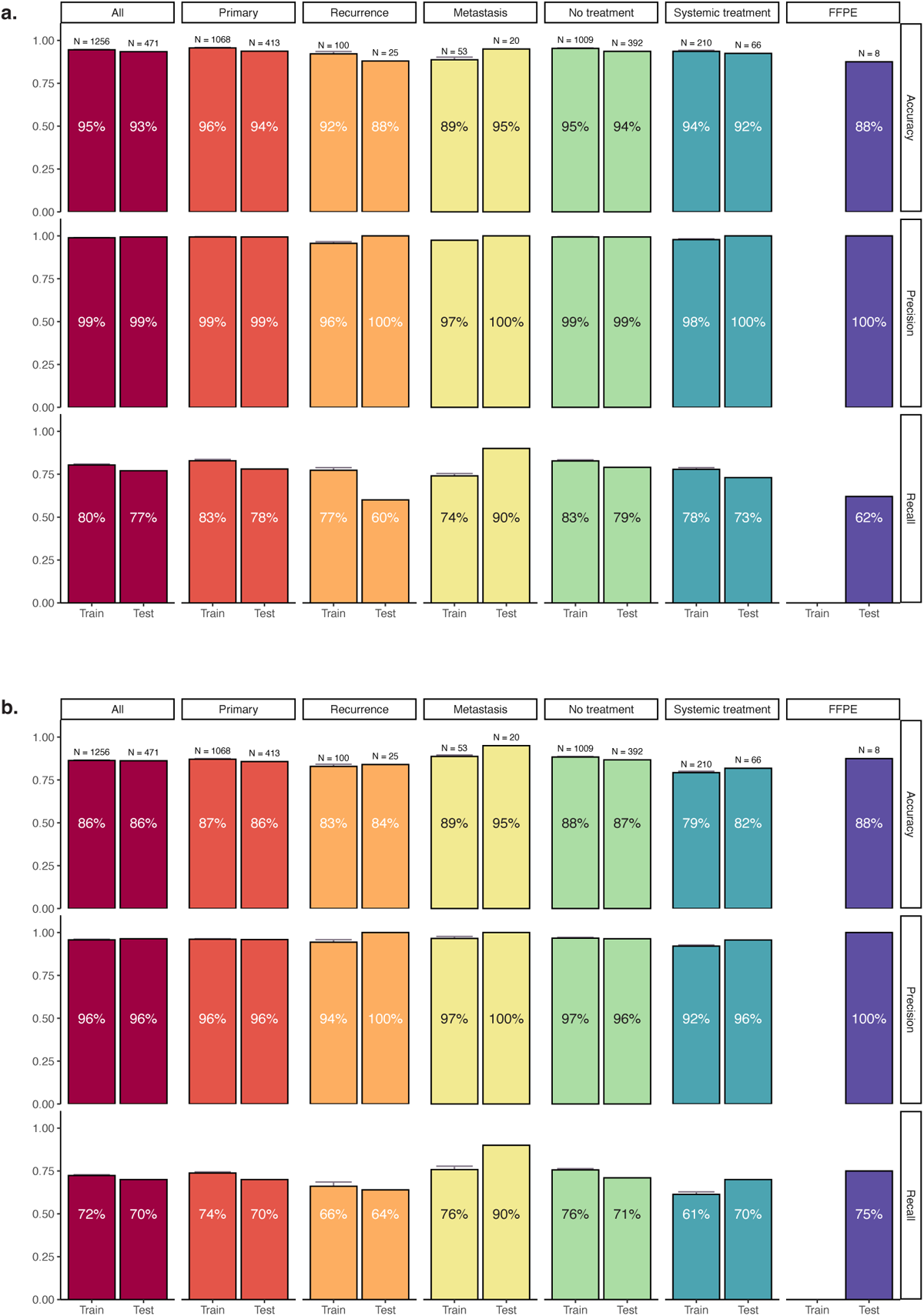
Performance of M&M on different sample types. Accuracy, precision and recall for the complete set of tumor type **(a)** and subtype **(b)** classifications grouped by their tumor and treatment status within the reference cohort (Train) and independent test cohort (Test).

---

**Supplementary Table 1. Overview of links between domain, tumor types, subtypes, abbreviations and the accompanying sample size. Available for download separately.**

**Supplementary Table 2.** Overview of M&M’s output on tumor type level. If there are less than three different labels administered within all models, the remaining predict and probability groups will be filled out with NA.

|  | predict | probability1 | predict2 | probability2 | predict3 | probability3 |
| --- | --- | --- | --- | --- | --- | --- |
| Patient1 | Pilocytic astrocytoma | 0.980 | Ganglioglioma | 0.003 | Paediatric-type diffuse low-grade glioma | 0.001 |
| Patient2 | Medulloblastoma | 1.000 | NA | NA | NA | NA |
| Patient3 | AML | 0.515 | Not malignant (Hemato) | 0.094 | JMML | 0.003 |

**Supplementary Table 3.** Overview of M&M’s output on tumor subtype level. If there are less than three different labels administered within all models, the remaining predict and probability groups will be filled out with NA.

|  | predict | probability1 | predict2 | probability2 | predict3 | probability3 |
| --- | --- | --- | --- | --- | --- | --- |
| Patient1 | Pilocytic astrocytoma | 0.980 | Ganglioglioma | 0.003 | Diffuse low-grade glioma, MAPK pathway-altered | 0.001 |
| Patient2 | Medulloblastoma, SHH-activated | 1.000 | NA | NA | NA | NA |
| Patient3 | GATA1 mutated AML/TMD | 0.100 | Bone marrow failure | 0.087 | Not malignant bone marrow | 0.007 |

**Supplementary Table 4.** Misdiagnoses within M&M algorithm with their top 3 classifications and probability scores. Available for download separately.

**Supplementary Table 5.** P-values associated with the comparisons of the performance measures between M&M and ALLCatchR on B-ALL subtype classifications. A star (*) signifies a significant difference (p-value *<* 0.05).

| Performance measure | Dataset | M&M better (p-value) | ALLCatchR better (p-value) |
| --- | --- | --- | --- |
| Accuracy | Reference cohort | 0.71 | 0.67 |
| Accuracy | Test cohort | 0.96 | 0.09 |
| Precision | Reference cohort | 0.06 | 0.94 |
| Precision | Test cohort | 1.00 | 0.71 |
| Recall | Reference cohort | 1.00 | 0.00* |
| Recall | Test cohort | 1.00 | 0.01* |

**Supplementary Table 6.**
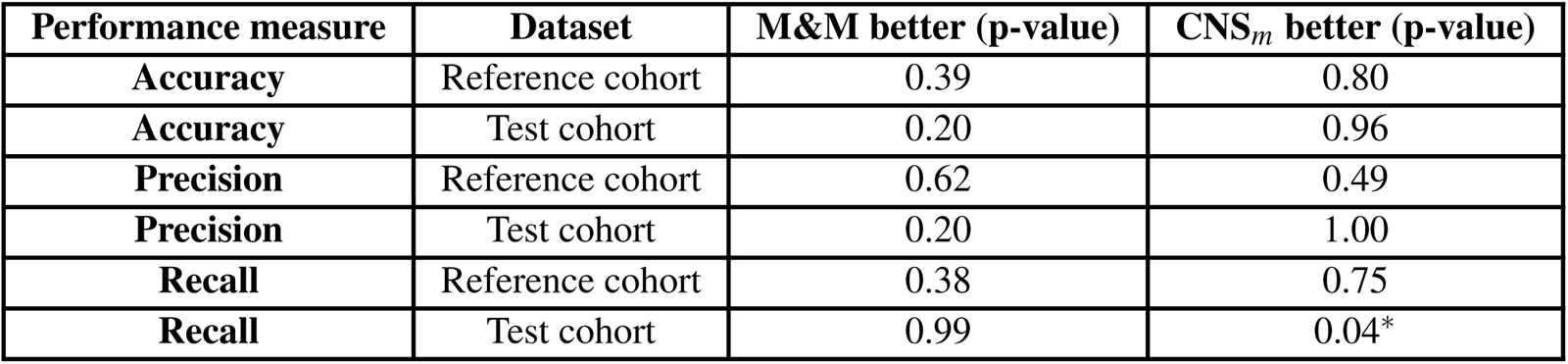
P-values associated with the comparisons of the performance measures between M&M and the DKFZ CNS methylation classifier (CNS*m*) on central nervous system tumor classifications. A star (*) signifies a significant difference (p-value *<* 0.05).

| Performance measure | Dataset | M&M better (p-value) | CNS<sub>m</sub> better (p-value) |
| --- | --- | --- | --- |
| Accuracy | Reference cohort | 0.39 | 0.80 |
| Accuracy | Test cohort | 0.20 | 0.96 |
| Precision | Reference cohort | 0.62 | 0.49 |
| Precision | Test cohort | 0.20 | 1.00 |
| Recall | Reference cohort | 0.38 | 0.75 |
| Recall | Test cohort | 0.99 | 0.04* |

**Supplementary Table 7.**
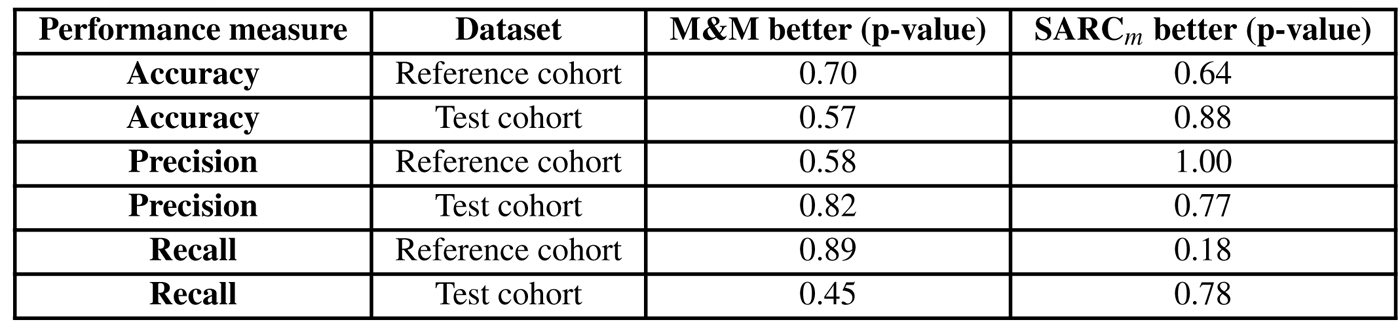
P-values associated with the comparisons of the performance measures between M&M and the DKFZ sarcoma methylation classifier (SARC*m*) on sarcoma classifications. A star (*) signifies a significant difference (p-value *<* 0.05).

**Supplementary Table 8. Tumor (sub)types that reach the sample size threshold of 3 upon combining the reference cohort and test cohort data, allowing their future inclusion into M&M**. Available for download separately.

